# Impact of Imaging Protocols on Thermal Detection of Pressure Injuries: Threshold versus Deep Learning Across Skin Tones

**DOI:** 10.64898/2026.05.21.26353842

**Authors:** Miriam Asare-Baiden, Sharon Eve Sonenblum, Kathleen Jordan, Glory Tomi John, Andrew Chung, Judy Wawira Gichoya, Vicki Stover Hertzberg, Joyce C. Ho

## Abstract

Pressure injuries represent a significant healthcare challenge requiring early detection to prevent severe complications. While thermal imaging shows promise for detecting early pressure related temperature changes, its robustness across varying imaging conditions and diverse patient populations remains unclear. This study systematically evaluated how imaging protocol variations (lighting, distance, positioning, camera type) and participant skin tone influence classification model performance for thermal cooling detection.

Using a controlled cooling protocol to simulate early pressure injury temperature changes, we collected 1,680 images from 35 diverse participants across 12 imaging protocol variations. We compared two approaches: three deep learning models (MobileNetV2, InceptionNetV3, ResNet50) and a threshold-based approach using an optimal fixed threshold temperature differential. Deep learning models outperformed the threshold-based approach, achieving 98.699.6% accuracy compared to 95.6%, with superior performance across all imaging protocols and skin tone groups. Threshold-based approach showed camera-dependent misclassification patterns across skin tones. On the high-resolution FLIR E8XT, the MST 7-10 group had 8 of 11 misclassifications. This pattern shifted on the low-resolution FLIR ONE Pro, where the intermediate skin tone group (MST 6) had 22 of 44 total misclassifications. In contrast, deep learning models maintained consistent performance across all skin tone groups and imaging protocols. Visualization analysis of the deep learning models suggested that these models focused on thermal gradients at cooling region boundaries, suggesting that spatial temperature gradients, not single-value thresholds, are critical for accurate detection. These findings suggest the potential of deep learning-based approaches to maintain robust, equitable performance across diverse skin tones and imaging conditions.

**Author Summary:** Pressure injuries are a major clinical challenge requiring early detection. Current visual inspection methods are unreliable, especially for patients with darker skin tones. Thermal imaging shows promise for detecting early temperature changes, but no studies have systematically evaluated how imaging variations affect detection accuracy across diverse populations. Using two cameras, FLIR E8XT (320×240 pixels) and FLIR ONE Pro (160×120 pixels), we collected 1,680 thermal images from 35 healthy adults across diverse skin tones within a controlled setting with simulated cooling and evaluated how imaging variations (lighting, distance, positioning, camera type) affect performance on two classification approaches: deep learning models that preserve spatial temporal context, and threshold-based approach using a single fixed temperature. Deep learning based approaches demonstrated superior robustness to camera type compared to the threshold based approach which exhibited hardware dependency, with performance deteriorating dramatically on the lower resolution FLIR ONE Pro camera. Deep learning-based approaches also showed consistent performance across all skin tone groups, indicating that both camera selection and labeling methodology are critical for clinical thermal imaging systems. Deep learning models preserve spatial temperature information and show promise for reliable pressure injury detection across diverse patient populations.

## Introduction

Pressure injuries are a major patient safety challenge with approximately 60,000 deaths occurring annually in the United States [1]. These localized injuries to the skin and underlying tissue result from prolonged pressure, shear, friction, or a combination of these mechanical factors, which usually develop over bony prominences or in areas where medical devices contact the skin [2]. Individual treatment costs vary dramatically based on severity, with hospital acquired pressure injury costs exceeding $26.8 billion annually in the United States [3]. Beyond direct treatment costs, pressure injuries contribute to extended hospital stays, increased nursing care requirements, legal liability, and reduced quality of life [4,5].

Early detection of pressure injuries is critical for preventing progression to severe stages, yet current assessment methods have significant limitations. Visual inspection [6] is a widely used approach that seeks to identify signs such as blanchable erythema, localized temperature changes, and participant reports of pain or unusual sensations in at-risk areas. However, visual inspection cannot reliably detect early signs of pressure injury development. Deep tissue changes, such as altered tissue consistency (firmness or sponginess), [2,7] often precede visible surface manifestations and non-blanchable erythema may be difficult to identify especially on darker skin tones [8]. Alternative technologies including subepidermal moisture measurement [9,10] and ultrasound [11] require direct contact with potentially damaged tissue and face cost barriers that prevent widespread implementation [12].

Thermal imaging, using long-wave infrared thermography, has shown promise in noncontact assessment of pressure injuries [13,14], aligning with clinical observations that temperature changes often precede skin color changes [15–17]. It can identify tissue ischemia across deep tissue injuries, from immobility-related injuries to device-related injuries, by detecting the thermal signatures of impaired blood flow before visible skin changes emerge [17,18]. Early detection protocols have demonstrated significant reductions in hospital-acquired pressure injury rates of up to 60% [13], enabling earlier intervention before visual identification is possible [19]. However, the clinical utility of thermal imaging depends critically on standardized image acquisition and interpretation protocols.

Previous research has definitively shown that in conventional optical photography, imaging conditions significantly affect the accuracy of pressure injury documentation, with variations in camera angle, distance, lighting, and image quality resulting in underestimation of wound area: camera angle changes alone can yield 35% measurement error [20,21]. Standardized protocols for controlling positioning, lighting, and camera parameters (angle, distance, rotation, height), along with trained research staff, are essential for effective wound imaging [22–24]. These principles often extend to thermal imaging application, though recent work suggests thermal imaging may be less sensitive to certain environmental factors such as lighting [25].

Beyond image acquisition standardization, using threshold-based decision-making to evaluate temperature differences presents its own challenges. This approach, common in clinical practice for translating quantitative measurements into actionable decisions [26], has been successfully applied in burn imaging where temperature difference >1°C predicted impaired healing [27]. Similarly, a Braden scale score of ≤ 18 is commonly used in clinical practice to identify patients at risk for pressure injury. However, one study identified the optimal cutoff to be 14 [17], suggesting that current thresholds may not reflect the best available evidence.

Establishing temperature thresholds for thermal imaging is complicated by technological variability. Thermal camera type significantly affects temperature measurements, with discrepancies exceeding 1°C between different devices limiting their interchangeability for establishing clinical detection thresholds [25]. This camera-dependent variation raises an important question: will temperature thresholds optimized on data from one camera generalize to images from different cameras or imaging conditions?

Deep learning offers a potential solution to these generalizability challenges by learning complex patterns from the data rather than relying on fixed temperature thresholds. Recent studies have developed deep learning models for pressure injury detection using thermal imaging [16,28], achieving mean average precision between 76.9% to 90.8% [29]. However, none have systematically evaluated how variations in imaging protocols affect model performance for pressure injury detection. These studies only focused on algorithm development under controlled conditions, without examining robustness to real-world imaging variations common in clinical settings. This is a critical evaluation gap given the potential advantage of the deep learning model over fixed thresholds. Additionally, none have evaluated performance across various skin tones or whether specific imaging protocols are needed for these deep learning models.

We build directly on Sonenblum et al.’s study [25], which established that camera type and skin tone affect the measured temperature change. We study whether these protocol variations affect the ability to classify thermal images (e.g., whether a given image represents cooling or not) by comparing two classification approaches, threshold-based detection and deep learning models, and evaluating their performance, reliability, and generalization across imaging protocols and skin tone groups.

## Methods

### Data Collection

This study leverages data from Sonenblum et al. [25], where data collection was designed as a controlled simulation to systematically evaluate how imaging protocol variations affect thermal imaging across diverse skin tones. The design used induced localized temperature changes in participants, which served as a controlled proxy for the early temperature changes associated with pressure injury development, where reduced blood flow to deformed tissue results in lower skin temperature before surface changes appear [30].

Thirty-five healthy adults were recruited between March to June 2024. Ethical approval for the study was obtained from the Emory University Institutional Review Board (eIRB number 00005999). Written informed consent was obtained from the participants prior to the data collection. All experiments were performed in accordance with the Emory University Institutional Review Board guidelines and regulations. Participants were characterized using the Monk Skin Tone (MST) scale [31], a 10-point classification system, by measuring the skin tone at the inner forearm. Recruitment initially targeted 30 participants with MST scores of 6 or higher to ensure adequate representation across darker skin tones, with an additional 5 participants with MST scores of 1-5 subsequently recruited to provide balance. For analytical purposes, participants were divided into three groups: MST 1-5 Group (n=5, 14.3%), MST 6 Group (n=7, 20%), and MST 7-10 Group (n=23, 65.7%). This stratification reflects evidence that MST 6 represents a critical transition point where pressure injury prevalence changes [32]. The distribution across MST levels and groups is shown in Table 1.

**Table 1.** Distribution of participants by MST scale. * Total images reflect acquisition across two devices

| Skin Tone Group | MST Level | Number of Participants (%) | Total Number of Images* |
| --- | --- | --- | --- |
| MST 1-5 Group | 1 | 0 (0.0) | 0 |
|  | 2 | 3 (8.6) | 144 |
|  | 3 | 0 (0.0) | 0 |
|  | 4 | 1 (2.9) | 48 |
|  | 5 | 1 (2.9) | 48 |
| MST 6 Group | 6 | 7 (20.0) | 336 |
| MST 7-10 Group | 7 | 22(62.9) | 1056 |
|  | 8 | 1 (2.9) | 48 |
|  | 9 | 0 (0.0) | 0 |
|  | 10 | 0 (0.0) | 0 |

Prior to image collection, two 2′ circles (left and right side) were marked on the posterior superior iliac spine (PSIS) of each participant. Temperature changes were induced at the right circle by placing a stone cylinder cooled to approximately 15.5°C via water bath on the right PSIS for 5 minutes. Images were captured using two cameras: the FLIR E8XT *(640 x 480 optical resolution, 320×240 thermal resolution)* and the FLIR ONE Pro *(1440 x 1080 optical resolution, 160×120 thermal resolution).* Image acquisition protocol followed a factorial design with two lighting conditions (ambient and ring light), two distances (35 and 50 cm), and three postures (forward placement of the top knee, stacked knees, backward placement of the top knee), resulting in 12 different imaging configurations. The order of these 12 configurations was randomly assigned and varied between participants. For each configuration and camera, we captured paired images before and after the cooling protocol, yielding a total of 48 images per participant (24 images per camera) and 1,680 images total (840 pre-cooling, 840 post-cooling). Pre-cooling and post-cooling images were labeled as baseline and cooling images, respectively. Fig 1 shows a series of baseline and cooling images taken under different imaging protocols.

**Fig 1.**
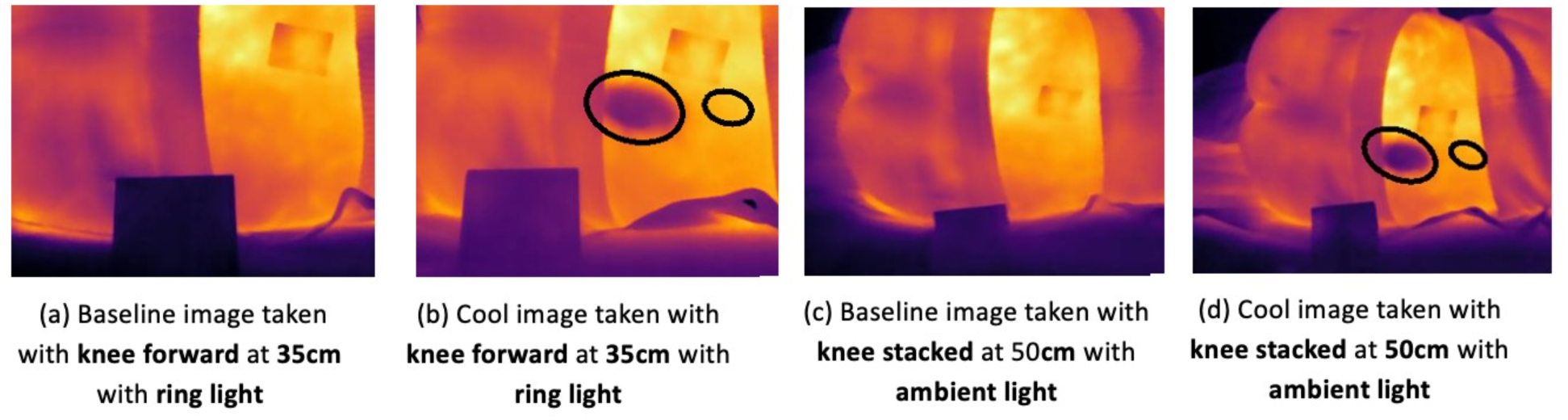
Baseline and cooling images taken at different lighting and posture for image protocols knee forward at 35 cm with ring light and knee stacked at 50 cm with ambient light, respectively. The big (left) ellipse represents the Region of Interest (ROI) and the smaller (right) ellipse represents the control region.

### Classification Approaches

Two approaches were evaluated for classifying thermal images as baseline or cooling. Threshold-based detection, the first approach, relies on identifying temperature differences relative to a healthy control region in the same image that exceeds a predetermined cutoff value. This approach has been successfully applied in burn imaging where temperature differences >1°C predicted impaired healing and is currently implemented clinically for early detection of pressure injuries, often with a threshold ranging anywhere between of 0.1°C [17,33], 1°C [34], 1.2°C [35], 1.5°C [36] and 2.2°C [37]. For this study, an optimal temperature differential threshold for the induced cooling was derived to classify images based on the temperature difference between the cooled region and the control region. While this is not the threshold used in clinical practice for pressure injury detection, this threshold is the data-driven cutoff optimized for distinguishing cooling-induced temperature changes from baseline conditions in our experimental protocol.

Convolutional neural networks (CNNs), a class of deep learning models specifically designed for image analysis, have become the standard approach for medical imaging tasks [38]. Popular CNN architectures including MobileNets [39], InceptionNets [40], and ResNets [41] are widely used for image analysis. These networks, initially trained on ImageNet’s 1.4 million images across 1,000 object categories, have achieved state-of-the-art performance in medical imaging through transfer learning. Transfer learning approach leverages knowledge from general object recognition (encoded by utilizing the pre-trained model weights) and applies it to specialized medical domains by fine-tuning the CNN to the domain-specific task [42]. This technique is particularly valuable as fine-tuning requires fewer samples given the limited availability of large, annotated clinical datasets.

CNNs have emerged as promising tools for pressure injury detection, with studies demonstrating the effectiveness of diverse architectures including VGG-16 and ResNet-50 for wound classification [43,44], encoder-decoder networks such as U-Net for precise lesion segmentation [45] and modern efficient architectures like MobileNets for pressure injury segmentation [28]. Given the limited dataset size (1,680 images, 35 participants), we prioritized established lightweight CNN architectures as more complex models do not yield performance improvements [46]. Three CNN architectures (MobileNetV2, ResNet50, InceptionNetV3) were evaluated for their ability to classify images as baseline or cooling across different imaging protocols.

### Classification Model Implementations

Images were locally normalized using per-image min-max scaling and resized using bilinear interpolation to match each model’s requirements: MobileNetV2 and ResNet50 (224×224 pixels) and InceptionNetV3 (299×299 pixels). To enhance model generalization and mitigate overfitting given the limited number of images, data augmentation was applied during finetuning. Specifically, we introduced horizontal flips, vertical flips, and rotation at 20° for each training image, which were empirically verified to improve model generalization on this dataset.

For all pre-trained architectures, the final classification layer was modified to output binary predictions (cooling or baseline). Fine-tuning followed conventional approaches used in transfer learning: Adam optimizer (learning rate: 0.001, batch size: 32) and binary cross-entropy loss. Training ran for a maximum of 100 epochs with early stopping to prevent overfitting.

For the threshold-based approach, an optimal temperature differential threshold of -1.71°C was derived using receiver operating characteristic (ROC) curve analysis on the full dataset. Images were classified as cooling if the temperature differential between the cooled region and the control region was ≤ -1.71°C, and as baseline otherwise. We note that unlike the CNN models, which were trained and evaluated on strictly disjoint folds, this threshold was optimized on the full dataset including test images and represents an optimistic estimate of threshold-based performance.

### CNN Explainability

To understand which image regions influenced the CNN predictions, Gradient-weighted Class Activation Mapping (Grad-CAM) was used to visualize the spatial features influencing classification decisions [47]. Grad-CAM highlights regions with the largest gradients in the final convolution layer, identifying areas most influential to cooling classification. This reveals whether the model focuses on clinically relevant anatomical features (the cooled region) or spurious correlations in the image.

### Evaluation setup

Model performance was assessed using a stratified, participant-level cross-validation framework. This approach maintained balanced skin tone group representation across all folds and prevented data leakage by ensuring individual participants’ images appeared exclusively in either training or testing sets within each fold. For the CNN models, fine-tuning was performed on four folds with evaluation on the held out fifth fold. For the threshold-based approach, the - 1.71°C threshold was applied uniformly to all images without training. Performance of both CNN and threshold-based approaches was assessed using four established classification metrics, accuracy, sensitivity, specificity, and F1 score, to provide a multidimensional evaluation of classification capability. Given the limited sample sizes in the MST 1-5 (n=5) and MST 6 (n=7) subgroups, statistical significance testing of cross-skin-tone performance differences was not conducted, as such tests would be underpowered to yield reliable conclusions.

## Results

### Overall Performance Comparison

Table 2 summarizes performance for the three CNN architectures and the threshold-based method for cooling detection. Overall values represent mean ± standard deviation from the 5-fold cross-validation.

**Table 2.** Performance for cooling detection across skin tone groups from the classification methods.

| Approach | Model | Skin Tone Group | Accuracy | F1 | Sensitivity | Specificity |
| --- | --- | --- | --- | --- | --- | --- |
| CNN-Based | MobileNetV2 | MST 1-5 | 0.975 | 0.974 | 0.950 | 1.000 |
|  |  | MST 6 | 0.994 | 0.994 | 0.988 | 1.000 |
|  |  | MST 7-10 | 0.993 | 0.993 | 0.986 | 1.000 |
|  |  | **Overall** | **0.990±0.009** | **0.990±0.010** | **0.981±0.019** | **1.000±0.000** |
|  | InceptionNetV3 | MST 1-5 | 0.983 | 0.983 | 0.975 | 0.992 |
|  |  | MST 6 | 1.000 | 1.000 | 1.000 | 1.000 |
|  |  | MST 7-10 | 0.997 | 0.997 | 0.998 | 0.996 |
|  |  | **Overall** | **0.996±0.006** | **0.996±0.006** | **0.995±0.007** | **0.996±0.005** |
|  | ResNet50 | MST 1-5 | 0.983 | 0.983 | 0.967 | 1.000 |
|  |  | MST 6 | 0.991 | 0.991 | 0.994 | 0.988 |
|  |  | MST 7-10 | 0.986 | 0.985 | 0.984 | 0.987 |
|  |  | **Overall** | **0.986±0.009** | **0.986±0.009** | **0.983±0.015** | **0.989±0.021** |
| Threshold-Based | - | MST 1-5 | 0.954 | 0.952 | 0.917 | 0.992 |
|  |  | MST 6 | 0.932 | 0.932 | 0.941 | 0.923 |
|  |  | MST 7-10 | 0.982 | 0.982 | 0.969 | 0.995 |
|  |  | **Overall** | **0.956** | **0.955** | **0.942** | **0.970** |

The CNN models consistently outperformed the threshold-based approach across all metrics. CNN accuracies ranged from 0.986 to 0.996, with InceptionNetV3 achieving the highest overall performance (accuracy = 0.996**±**0.006, F1 = 0.996**±**0.006), followed by MobileNetV2 (accuracy = 0.990**±**0.009, F1 = 0.990**±**0.010), and ResNet50 (accuracy = 0.986**±**0.009, F1 = 0.986**±**0.009). In contrast, the threshold-based approach using the ROC-optimized temperature differential threshold of -1.71°C achieved lower performance with an overall accuracy of 0.956 (F1 = 0.955).

### Performance Across Skin Tone Groups

The CNN models demonstrated slight variation across skin tone groups, with MST 6 Group and MST 7-10 Group generally yielding better performance compared to MST 1-5 Group. InceptionNetV3 achieved perfect performance for MST 6 Group on all four metrics, while MobileNetV2 yielded perfect specificity on all three groups. Even for MST 1-5 Group, the CNN models maintained excellent performance, with accuracies ranging from 0.975 (MobileNetV2) to 0.983 (InceptionNetV3 and ResNet50). Notably, all CNN models maintained perfect or near perfect specificity across skin tone groups, indicating excellent ability to correctly detect cooling regardless of skin tone.

Performance of the threshold-based approach varied by skin tone group, with MST 1-5 achieving 0.954 accuracy, MST 6 achieving 0.932, and MST 7-10 achieving 0.982 accuracy. Compared to CNN models, threshold-based accuracy, F1, and sensitivity were lower across all skin tones, with differences ranging from 0.015 to 0.068. The largest gap occurred in MST 6 (0.068 accuracy difference), representing approximately 21 additional misclassifications on the 336 images in this group compared to average CNN performance. This suggests that intermediate skin tones are particularly vulnerable to fixed threshold classification, as using a fixed temperature threshold may not equally capture cooling signatures across different skin tone populations.

### Protocol-Specific Performance Analysis

We performed detailed protocol-level analysis using MobileNetV2 as it combines lightweight computational requirements with strong performance characteristics. Its position between InceptionNetV3 (highest performance) and ResNet50 (lowest performance), as shown in Table 2, makes it a representative model for evaluating how imaging protocols affect CNN-based cooling detection across diverse settings.

Table 3 summarizes the effect of each imaging protocol on the two classification methods: MobileNetV2 and threshold-based approach.

**Table 3.** Analysis of the number of misclassified images based on protocol.

| Protocol | Posture | Lighting | Distance<br>(cm) | MobileNetV2 |  | Threshold |  |
| --- | --- | --- | --- | --- | --- | --- | --- |
|  |  |  |  | Count | (%) | Count | (%) |
| 1 | Forward | Ambient | 35 | 2 | (2.9) | 4 | (7.0) |
| 2 | Forward | Ambient | 50 | 2 | (2.9) | 7 | (13.0) |
| 3 | Forward | Ring | 35 | 4 | (5.7) | 7 | (13.0) |
| 4 | Forward | Ring | 50 | 2 | (2.9) | 6 | (11.6) |
| 5 | Stacked | Ambient | 35 | 2 | (2.9) | 5 | (9.3) |
| 6 | Stacked | Ambient | 50 | 0 | (0.0) | 3 | (5.6) |
| 7 | Stacked | Ring | 35 | 0 | (0.0) | 5 | (9.3) |
| 8 | Stacked | Ring | 50 | 1 | (1.4) | 5 | (9.3) |
| 9 | Back | Ambient | 35 | 0 | (0.0) | 4 | (7.4) |
| 10 | Back | Ambient | 50 | 0 | (0.0) | 1 | (1.9) |
| 11 | Back | Ring | 35 | 2 | (2.9) | 3 | (5.6) |
| 12 | Back | Ring | 50 | 1 | (1.4) | 4 | (7.4) |
| **Total** |  |  |  | **16** | **(1.0)** | **54** | **(3.2)** |

MobileNetV2 produced a total of 16 misclassifications across the 12 image acquisition protocols. Protocol 3 (knee forward, ring light, 35cm distance) had the highest error count with misclassifications (5.7%), while protocols 6 (knee stacked, ambient, 50 cm distance), 7 (knee stacked, ring, 35 cm), 9 (knee back, ambient, 35 cm), and 10 (knee back, ambient, 50 cm distance) achieved perfect classification (0 misclassifications). The remaining protocols had 1-2 misclassifications each (1.4-2.9%) indicating robust performance across imaging conditions despite minor protocol-specific variations.

The threshold-based approach, however, produced 54 total misclassifications across the 12 protocols (3 times more errors than MobileNetV2). Protocols 2 (knee forward, ambient, 50 cm) and 3 (knee forward, ring, 35 cm) had the highest error counts with 7 misclassifications each (13.0 %), while Protocol 10 (knee back, ambient, 50 cm) had only one misclassification (1.9%). Misclassifications across the remaining protocols ranged from 3 to 6 (5.6% to 11.6%), with errors distributed relatively evenly across both ring light (Protocols 3, 4, 7, 8, 11, 12) and ambient light conditions. Overall, ambient light had fewer misclassifications in comparison to ring light across the protocols (44.4% vs. 55.6%).

#### Performance by Camera Type

We further examined misclassification patterns across the two camera types (FLIR E8XT and FLIR ONE Pro) for both MobileNetV2 and the threshold-based approach. This analysis addresses whether MobileNetV2, which preserves temporal and spatial context, provides advantages over threshold-based classification derived from optimized temperature delta (ΔT) analysis across the two cameras and skin tone groups as shown in Table 4.

**Table 4.** Misclassified images by camera type based on skin tone.

| Skin Tone Group | FLIR E8XT |  |  |  | FLIR One Pro |  |  |  |
| --- | --- | --- | --- | --- | --- | --- | --- | --- |
|  | MobileNetV2 |  | Threshold |  | MobileNetV2 |  | Threshold |  |
|  | Count | (%) | Count | (%) | Count | (%) | Count | (%) |
| 1-5 | 4 | (3.3) | 1 | (0.8) | 2 | (1.7) | 10 | (8.3) |
| 6 | 1 | (0.6) | 1 | (0.6) | 1 | (0.6) | 22 | (13.1) |
| 7-10 | 4 | (0.7) | 8 | (1.5) | 4 | (0.7) | 12 | (2.2) |
| **Total** | **9** | **(1.1)** | **10** | **(1.2)** | **7** | **(0.8)** | **44** | **(5.2)** |

##### FLIR E8XT Performance

MobileNetV2 and threshold-based approaches demonstrated lower overall misclassification rates (1.1% and 1.2%, respectively). The highest error rates on MobileNetV2 occurred on the lighter and darker skin tone (MST 1-5), with each having 4 misclassifications. However, on the threshold-based approach, misclassifications were highest on the darker skin tones (8 out of 10). While MobileNetV2 had four (3.3%) misclassifications on the lighter skin tone, only one (0.8%) image was misclassified on the threshold-based approach.

#### FLIR ONE Pro Performance

The threshold-based approach demonstrated substantially higher misclassification rates (5.2%) compared to MobileNetV2 (0.8%), representing an 84% reduction in errors. The threshold-based approach showed elevated error rates across all skin tone groups, with 22 times more errors in MST 6 group compared with the FLIR E8XT camera (13.1% vs. 0.6%). In contrast, MobileNetV2 recorded the highest error rate in the lightest skin tone, MST 1-5 (1.7%), which was 5 times lower than threshold-based approach on the same camera. Notably, the error rates across the three skin tone groups were substantially lower on MobileNetV2 with the FLIR E8XT.

#### Error Analysis on MobileNetV2 Misclassifications

To understand the sources of misclassification, we conducted detailed error analyses using Grad-CAM visualizations on two representative cases: one from the MST 1-5 Group and one from the MST 7-10 Group. Grad-CAM heatmaps overlay attention maps on the thermal images with red regions indicating areas where MobileNetV2 focused its attention during classification and blue regions representing less important areas.

Although three CNN architectures were evaluated with InceptionNetV3 achieving the highest overall accuracy (Table 2), we focus on MobileNetV2 misclassifications for detailed error analysis because they exemplify typical failure patterns observed across all three models, providing comprehensive insight into the failure points that can affect CNN-based thermal cooling detection. The following analysis examines how temporal changes in thermal distributions and skin tones impact CNN attention patterns and classification outcomes. Both cases were captured using the FLIR E8XT camera, which exhibits greater temperature sensitivity and thermal contrast compared to the FLIR ONE Pro [25]. This enhanced thermal range makes it suitable for identifying subtle temperature variations that could influence model classification behavior. The thermal images use standard temperature-to-color mapping, where warmer temperatures correspond to brighter colors (white, yellow, orange) and cooler temperatures to darker colors (purple, blue, black). Note that baseline skin temperatures differ between the selected participants, with Fig 2 showing higher overall temperatures than Fig 3.

**Fig 2.**
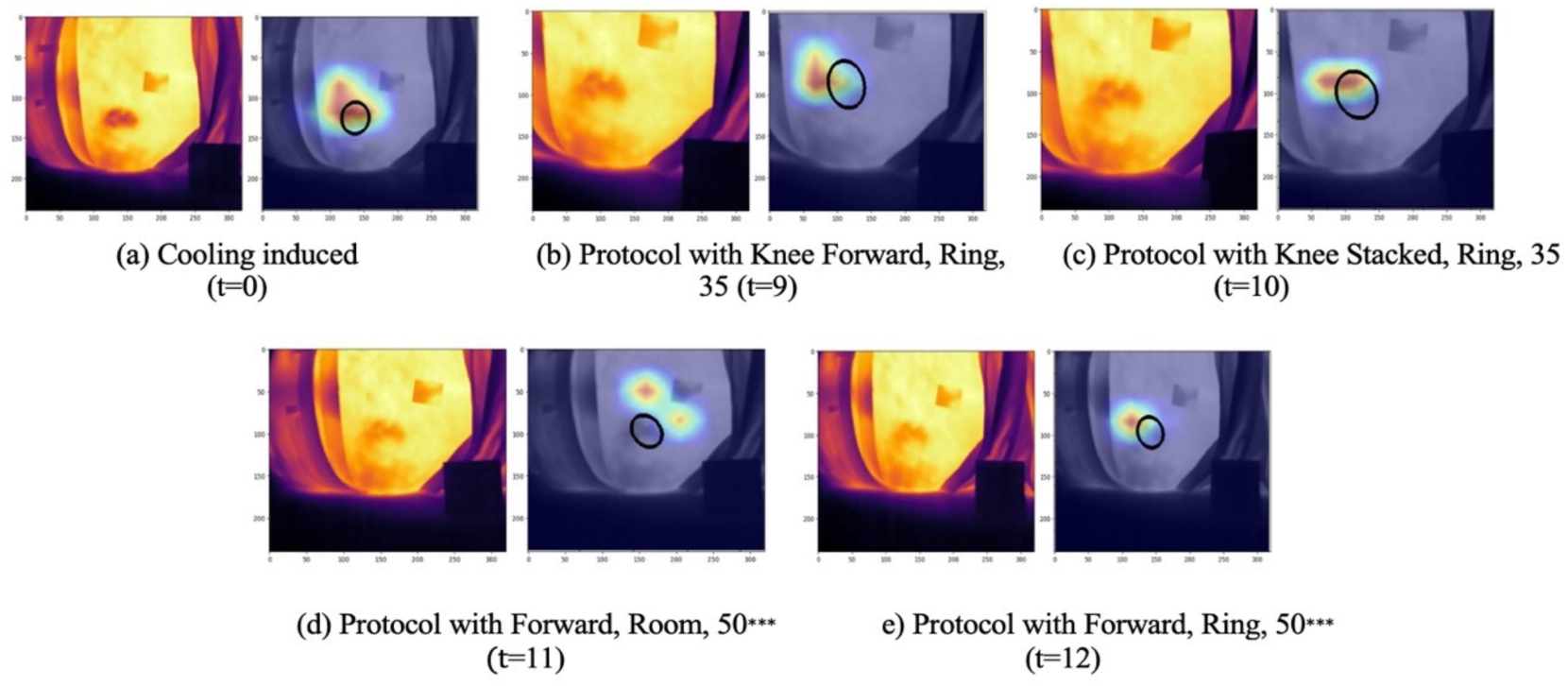
The progression of thermal images for the cooling protocol of a MST 7-10 Group participant and their associated Grad-CAM visualization. For each set of thermal images, the left displays the original image and the right overlays the region with the largest impact on the prediction. The number in parenthesis (t=x) indicates the image acquisition sequence number. *** represents the misclassified images and black circles represents the region of interest.

**Fig 3.**
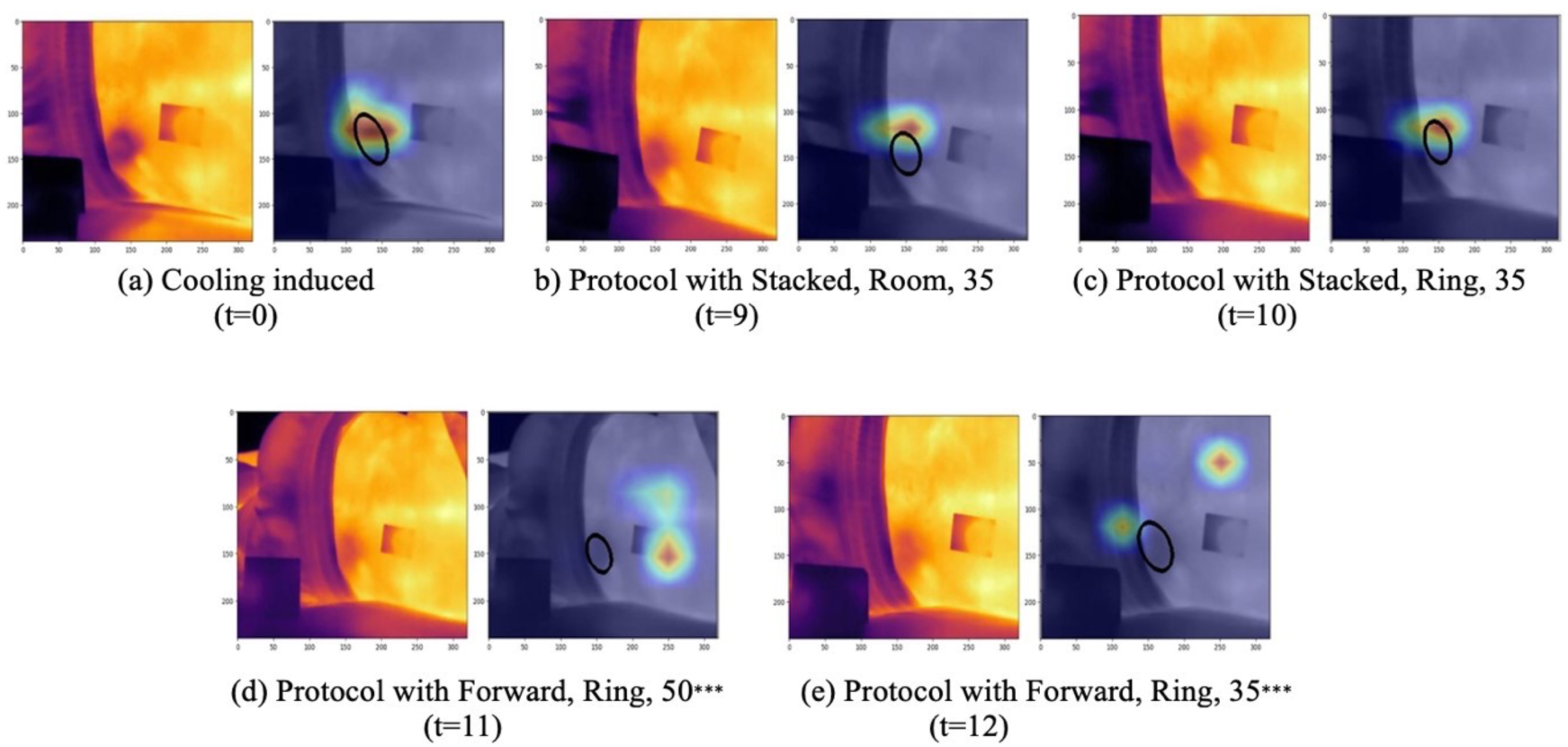
The progression of thermal images for the cooling protocol of a MST 1-5 Group participant and their associated Grad-CAM visualization. For each set of thermal images, the left displays the original image and the right overlays the region with the largest impact on the prediction. The number in parenthesis (t=x) indicates the image acquisition sequence number. *** represents the misclassified images and black circles represents the region of interest.

##### MST 7-10 Group Participant Analysis

Fig 2 presents the image collection sequence for a participant in the MST 7-10 Group, illustrating the evolution of thermal patterns following the cooling protocol. The first post-cooling image (Fig 2(a)) shows uniform cooler temperatures across the participant’s right posterior superior iliac spine region. The corresponding Grad-CAM visualization (right image of Fig 2(a)) confirms that MobileNetV2 correctly focused on the cooled region itself, with concentrated red attention patterns in the center of the cooling area. As the image acquisition progressed from t=9 to t=12 (Fig s 2(b)-2(e)), the model’s behavior shifted in response to thermal changes within the cooling region.

Temperature differential measurements (ΔT) quantify this thermal evolution. Immediately post-cooling (Fig 2(a)), the cooling region showed a pronounced temperature differential of 4.12°C. However, by the ninth timepoint (Fig 2(b)), this had warmed substantially to -1.92°C, and the rewarming continued through the final acquisition, reaching -1.82°C by the twelfth timepoint (Fig 2(e)). This rapid 2.3°C recovery reduced the thermal contrast that MobileNetV2 had learned to recognize, leading to misclassifications.

Correspondingly, Grad-CAM visualizations demonstrate how the model’s attention followed these thermal dynamics: the model correctly focused on the temperature gradient at the cooling region boundary when thermal contrast was pronounced (Fig s 2(b)-2(c)), with concentrated red attention patterns. However, at t=11 (Fig 2(d)), when the cooling region’s thermal contrast had diminished substantially, MobileNetV2’s focus shifted away from the cooling region boundary toward localized warming artifacts that had become increasingly pronounced within the cooling region. By t=12 (Fig 2(e)), the Grad-CAM heatmap shows a small, localized blue attention pattern, indicating the model struggled to identify clear cooling signals with minimal thermal gradient (ΔT = -1.82°C) remaining at that timepoint.

##### MST 1-5 Group Participant Analysis

The image collection sequence after the cooling protocol for a participant from MST 1-5 Group is shown in Fig 3. Similar to the MST 7-10 Group participant, the first post-cooling image (Fig 3(a)) exhibits the uniform cooling region on the participant’s right posterior superior iliac spine region. Similarly, the last two image acquisitions (Fig s 3(d) and 3(e)) were incorrectly classified. However, the thermal dynamics and corresponding Grad-CAM patterns reveal a different mechanism. Cooling region rewarming progressed gradually with an initial ΔT of - 3.78°C immediately post-cooling (Fig 3(a)), warming to -2.73°C by the ninth timepoint (Fig 3(b)) and reaching -2.34°C by the final acquisition (Fig 3(e)). Despite this gradual rewarming, the cooling region maintained substantial thermal contrast throughout the acquisition sequence (ΔT remained well below -2.3°C at all timepoints).

Yet despite this maintained thermal contrast, the participant experienced misclassifications at later timepoints (Fig s 3(d)-3(e)). Grad-CAM analysis reveals that the model’s attention shifted not toward the diminishing cooling region boundary (as in the MST 710 participant), but toward pronounced paraspinal warming in adjacent anatomical regions. Areas closer to the spine and the left paraspinal region exhibited increased skin temperatures during the sequence, creating competing thermal gradients that captured the model’s attention. This is a critical distinction: the ΔT measurements confirm the cooling region remained cold and thermally distinct, yet the model still misclassified because its learned attention mechanisms responded to competing warm regions with stronger local thermal contrast.

Interestingly, the correctly classified image at t=10 (Fig 3(c)) differed from the misclassified image at t=11 (Fig 3(e)) only in collection protocol (knee position: stacked versus forward). This minimal protocol difference suggests that simultaneous rewarming of the cooling region and intensifying paraspinal warming, creating patterns that the model could not reliably distinguish, rather than the postural variation, are the primary factors driving misclassification in this participant.

#### Cross-Participant Pattern Identification

Comparative analysis of both participants revealed consistent error patterns across the two skin tone groups. In both cases, MobileNetV2 correctly classified images when thermal gradients at the cooling region boundary were strong early in the image acquisition sequence, but the few misclassifications occurred as these thermal gradients reduced over time. Grad-CAM visualizations demonstrated that over time, the model’s attention is impacted by warmer and higher contrast regions rather than remaining focused on the cooling region boundary with its reduced thermal gradient. Despite different anatomical locations of warming (paraspinal regions in the MST 1-5 Group participant versus warm spots on or near the cooling region in the MST 710 Group participant), the attention-shifting behavior towards stronger thermal gradients were consistent across both cases, suggesting participant-independent model behavior rather than individual participant characteristics or skin tone differences.

## Discussion

This study systematically compared two classification approaches for thermal imaging-based cooling detection: CNN models and threshold-based approach using an ROC-optimized temperature differential of -1.71°C. CNN models outperformed the threshold-based approach, achieving 98.6-99.6% accuracy compared to 95.6%, with performance advantages consistent across imaging protocols and skin tone groups.

The improved performance of the CNN models reflects a fundamental difference in how these approaches capture thermal information. The threshold-based approach reduces complex thermal images to a single temperature differential value, discarding spatial information about where temperature changes occur and how thermal gradients are distributed across the region of interest. In contrast, CNN models preserve spatial patterns, learning to recognize thermal gradients and their relationship to cooling-induced changes. This is particularly important for pressure injury detection, where spatial temperature distribution, not just a single temperature measurement, characterizes early tissue injury. Furthermore, selection of an appropriate control region for a threshold-based approach may be especially subjective in clinical application, as it depends on clinician judgment and anatomical and thermal variability. Grad-CAM analysis confirmed that MobileNetV2 learned to focus on thermal gradients at cooling region boundaries, demonstrating attention to clinically relevant spatial features rather than spurious correlations.

Performance variations across imaging protocols further illuminate this distinction. The threshold-based approach showed protocol-dependent failures, with error rates ranging from 1.9% (Protocol 10) to 13.0% (Protocols 2, 3), and ring light misclassifying more images (30 total) compared to ambient light (24 images). In contrast, MobileNetV2 maintained consistent performance across these same conditions, with four protocols (6, 7, 9, and 10) achieving zero misclassifications and error rates distributed without systematic patterns. However, future studies with larger datasets are needed to confirm whether these protocol-dependent patterns persist.

The two approaches showed different sensitivity to camera type and hardware. On the FLIR E8XT, performance was broadly comparable overall, though threshold-based methods outperformed MobileNetV2 for lighter skin tones, suggesting MobileNetV2 may have learned features less discriminative for lighter skin tones in high-resolution thermal data. On the consumer-grade FLIR One Pro, the performance gap widened dramatically, with threshold failures concentrated in intermediate skin tones. This hardware dependency of threshold-based approaches reflects their reliance on specific thermal signatures produced by different sensor designs, whereas CNN models learned spatial features generalize better across camera types. The skin tone-dependent performance suggests that physiological factors such as melanin content, subcutaneous tissue composition, or microvascular density may influence thermal emission or heat dissipation patterns in ways a fixed temperature thresholds cannot accommodate, particularly when thermal resolution is limited. These findings suggest that the measurement level effects documented by Sonenblum et al. [25], extend to classification performance, with camera type and skin tone influencing not only thermal measurements but also how reliably CNN models and threshold-based methods can distinguish cooling from baseline images. Notably, that study found higher melanin content produced *larger* temperature differentials after cooling, which might suggest darker skin tones would be easier to classify. Threshold-based methods showed camera-dependent vulnerability patterns. On the high-resolution E8XT, darker skin tones (MST 7-10) had the highest misclassification rates (8 misclassifications in total). However, on the low-resolution ONE Pro, the vulnerability shifted to intermediate skin tones (MST 6) which had a total of 22 misclassifications. This resolution-dependent shift in failure patterns suggests that thermal resolution and skin tone physiology interact in complex ways that fixed temperature thresholds cannot accommodate. CNN models, by learning spatial gradient patterns rather than relying on a single temperature value, appeared less vulnerable to this compression effect and maintained robust and equitable performance across cameras and skin tone groups.

Grad-CAM analysis of representative MobileNetV2 misclassifications suggests a failure mode related to temporal rewarming effects. In both cases examined, MobileNetV2 correctly classified early images when thermal gradients at the cooling region boundaries were pronounced but misclassified later images as cooling regions began to rewarm. Grad-CAM visualizations showed that model attention shifted away from diminishing cooling region boundaries toward emerging localized warm spots (paraspinal regions or adjacent warming areas) that emerged as thermal gradients diminished. This attention-shifting behavior was consistent across both cases examined and suggests that MobileNetV2’s misclassifications stem from thermal pattern ambiguity, rather than skin-tone-specific differences. Both classification approaches face this temporal challenge. For threshold-based methods, progressive temperature decay eventually crosses the fixed threshold, while for CNN models, diminishing boundary gradients reduce discriminative features while competing warm regions capture model attention. CNN models’ lower overall error rates nevertheless suggest their spatial pattern recognition provides greater resilience against these temporal confounders.

These findings have important implications for thermal imaging-based pressure injury detection. While fixed temperature thresholds of 0.1°C [17,33], 1°C [34], 1.2°C [35], 1.5°C [36] and 2.2°C [37] are conceptually appealing for clinical implementation, they discard spatial information and cannot adapt to the temporal dynamics of thermal recovery that characterize real physiological responses. However, robust clinical deployment of CNN-based systems requires training datasets that capture complete thermal recovery trajectories and diverse anatomical variations. As temperature differentials approach clinical thresholds, single-frame classification may become unreliable, particularly in anatomically variable regions such as the gluteal cleft. Temporal sequences with precise labeling could address this by capturing thermal trajectories rather than isolated temperature values. Nevertheless, nurse oversight remains essential to contextualize model outputs and differentiate clinically meaningful thermal anomalies from physiological ones.

For clinical translation, these findings suggest that dataset development should prioritize labeling based on known clinical conditions (e.g., confirmed pressure injury vs healthy tissue) rather than on post-hoc temperature threshold criteria., CNN models consistently high performance across the 12 tested protocols indicates clinical implementation does not require rigid standardization of imaging conditions. CNN models can tolerate natural variations in distance, lighting, and patient positioning that inevitably occur in real-world clinical settings. Although certain protocols 6, 7, 9, and 10 achieved zero misclassifications and may represent optimal image acquisition conditions, the overall robustness of CNN models suggests that clinical users can focus on capturing clear thermal images without being constrained by strict protocol requirements.

Several limitations should be considered when interpreting our findings. First, localized external cooling was used as a proxy for ischemia-driven pressure injury temperature changes rather than actual pressure injury development. While Sonenblum et al. [25] validated that this protocol produces robust, clinically meaningful temperature differentials detectable across all skin tones, external cooling and ischemic cooling differ in their underlying mechanisms and may produce thermal gradients with different spatial morphologies. How the difference affects CNN model generalization to clinical ischemia signatures remains unknown. Additionally, pressure injury develops gradually over hours or days, while our protocol captured acute cooling and rewarming over a single session, which may limit the generalizability to clinical scenarios.

Second, the healthy adult cohort may not reflect clinical populations with comorbidities and physiological variations that could influence thermal imaging performance. Third, our recruitment strategy prioritized darker skin tones (MST ≥6), resulting in limited sample sizes for the MST 1-5 and MST 6 Groups. These small subgroups limit statistical power for detecting protocol-specific performance differences in these groups. Findings for these groups, including the threshold-based approach’s elevated error rate in MST 6, should be interpreted with caution. Finally, images were captured over a limited time window following cooling; longitudinal assessment of thermal pattern evolution in developing pressure injuries is needed.

Future validation in clinical populations with actual pressure injury risk and larger balanced cohorts across all skin tone groups are essential to confirm the clinical utility of CNN based thermal imaging detection. Prospective studies should evaluate whether the improved equity and robustness of CNN models observed in this controlled setting translate to improved early detection of pressure injuries in real clinical environments, where additional challenges such as patient movement, varying ambient temperatures, and time constraints may affect performance.

Beyond technical validation, future research should address two critical concerns about AI-based thermal imaging deployment. First, systematic evaluation is needed to ensure CNN models learn clinically meaningful thermal patterns rather than spurious correlations with imaging artifacts, equipment characteristics, or other non-physiological features that may inadvertently correlate with patient demographics [48]. Our Grad-CAM analysis demonstrated attention to thermal gradients at cooling boundaries, but similar validation in clinical datasets is essential to confirm models focus on tissue-level thermal changes indicative of pressure injury rather than confounding factors. Sonenblum et al. [25] found melanin content correlates with cooling response magnitude, but whether this reflects physiology or measurement artifacts is unclear. Models trained on such data may learn demographic-correlated artifacts as features, underscoring the need for explainability validation across populations. Second, even when models demonstrate technical equity across skin tones, research should assess whether clinicians differentially trust or scrutinize AI-generated assessments based on patient characteristics [49,50]. Understanding both what CNN models learn and how clinicians integrate these assessments with clinical judgement will be crucial for ensuring equitable pressure injury detection across diverse patient population.

## Conclusion

CNN-based models outperformed threshold-based approaches for detecting cooling-induced thermal signatures across diverse imaging protocols and skin tones. This performance advantage was consistent across both camera types and all skin tone groups, with CNN models demonstrating particular robustness on the lower-resolution FLIR ONE Pro camera. Grad-CAM analysis revealed that CNN models learned to focus on thermal gradients at cooling region boundaries which are clinically relevant spatial features that threshold-based approaches cannot capture.

For clinical translation, these findings suggest that thermal imaging datasets for pressure injury detection should be labeled based on clinical ground truth (confirmed injury status) rather than post-hoc temperature thresholds. CNN models’ robustness across imaging protocols indicates clinical implementation does not require rigid standardization, allowing natural variations in distance, lighting, and positioning that inevitably occur in real-world settings. Future validation in at-risk clinical populations with larger balanced cohorts across skin tones is essential to confirm whether the technical equity and robustness observed here translate to improved early detection and equitable care.

## Data Availability

The minimal dataset and accompanying code is available at Google Drive via https://drive.google.com/drive/folders/1UuthAg6h1uWE4LuBveFyx68GZCCOFAri?usp=sharing Upon acceptance dataset and code will be made publicly available via Zenodo and GitHub respectively

https://doi.org/10.5281/zenodo.16923157.

## Acknowledgments

This work was supported in part by the National Center for Advancing Translational Sciences of the National Institutes of Health under Award number UL1TR002378 and the National Institute of Biomedical Imaging and Bioengineering of the National Institutes of Health under Award Number R01EB036579. The content is solely the responsibility of the authors and does not necessarily represent the official views of the National Institutes of Health.

